# Association Between Age And Endovascular Treatment Outcomes: Binational Registry Of 9941 EVT Cases From Korea And Taiwan

**DOI:** 10.1101/2024.12.31.24319838

**Authors:** Beom Joon Kim, Sung-Chun Tang, Yi-Chen Hsieh, Chih-Hao Chen, Yong Soo Kim, Chun-Jen Lin, Jong-Moo Park, Pi-Shan Sung, Kyusik Kang, Yu-Wei Chen, Soo Joo Lee, Kuan-Hung Lin, Jae-Kwan Cha, Chih-Wei Tang, Tai Hwan Park, Hai-Jui Chu, Kyungbok Lee, Chuan-Hsiu Fu, Jun Lee, Chao-Liang Chou, Keun-Sik Hong, Ching-Huang Lin, Kyung-Ho Yu, Cheng-Yu Wei, Dong-Eog Kim, Shang-Yih Yen, Joon-Tae Kim, Po-Lin Chen, Jay Chol Choi, Lung Chan, Jee Hyun Kwon, Dong-Ick Shin, Sung-Il Sohn, Hung-Yi Chiou, Chulho Kim, Kwang-Yeol Park, Chi Kyung Kim, Li-Ming Lien, Sung Hyuk Heo, Jiunn-Tay Lee, Hee-Joon Bae, Jiann-Shing Jeng

**Affiliations:** Department of Neurology, Seoul National University Bundang Hospital, Seongnam-si, Gyeonggi-do, Republic of Korea; Department of Neurology, National Taiwan University Hospital, Taiwan; Program in Medical Neuroscience, Taipei Medical University, Taiwan; Department of Neurology, Neurological Institute, Taipei Veterans General Hospital, Taiwan; Department of Neurology, Uijeongbu Eulji Medical center, Eulji University, Uijenongbu, Korea; Department of Neurology, National Cheng Kung University Hospital, College of Medicine, National Cheng Kung University, Tainan, Taiwan; Department of Neurology, Nowon Eulji Medical Center, Eulji University School of Medicine, Seoul, Korea; Department of Neurology, Landseed International Hospital, Taoyuan, Taiwan; Department of Neurology, Eulji University, School of Medicine, Daejeon Eulji Medical Center, Daejeon, Korea; Department of Neurology, Chi Mei Medical Center, Tainan, Taiwan; Department of Neurology, Dong-A University Hospital, Busan, Korea; Department of Neurology, Far Eastern Memorial Hospital, New Taipei City, Taiwan; Department of Neurology, Seoul Medical Center, Seoul, Korea; Department of Neurology, En Chu Kong Hospital, New Taipei City, Taiwan; Department of Neurology, Soonchunhyang University Seoul Hospital, Seoul, Korea; Department of Neurology, National Taiwan University Hospital Hsin-Chu Branch, Hsinchu City; Department of Neurology, Yeungnam University Medical Center, Daegu, Korea; Department of Neurology, Mackay Memorial Hospital, Taipei, Taiwan; Department of Neurology, Inje University Ilsan Paik Hospital, Goyang, Korea; Department of Neurology, Kaohsiung Veterans General Hospital, Taiwan; Department of Neurology, Hallym University Sacred Heart Hospital, Anyang, Korea; Department of Neurology, Chang Bing Show Chwan Memorial Hospital, Changhwa County, Taiwan; Department of Neurology, Dongguk University Ilsan Hospital, Goyang, Korea; Department of Neurology, Tri-Service General Hospital, National Defense Medical Center, Taipei, Taiwan; Department of Neurology, Chonnam National University Medical School, Chonnam National University Hospital, Gwangju, Korea; Department of Neurology, Taichung Veterans General Hospital, Taiwan; Department of Neurology, Jeju National University Hospital, Jeju, Korea; Department of Neurology, Taipei Medical University-Shuang Ho Hospital, New Taipei City, Taiwan; Department of Neurology, Ulsan University Hospital, University of Ulsan College of Medicine, Ulsan, Korea; Chungbuk National University Hospital, Chungbuk National University College of Medicine, Cheongju, Korea; Department of Neurology, Keimyung University Dongsan hospital, Keimyung University School of Medicine, Daegu, Korea; School of Public Health, College of Public Health, Taipei Medical University, Taiwan; Department of Neurology, Hallym University Chuncheon Sacred Heart Hospital, Gangwon-do, Korea; Department of Neurology, Chung-Ang University Hospital, Seoul, Korea; Department of Neurology, Korea University College of Medicine, Seoul, Korea.; Department of Neurology, Korea University Guro Hospital, Seoul, Korea; Department of Neurology, Shin Kong WHS Memorial Hospital, Taipei, Taiwan; Department of Neurology, Kyung Hee University Hospital, Seoul, Korea; Department of Neuroscience, College of Medicine, Seoul National University, Seoul, Republic of Korea

## Abstract

**Background and Purpose:** As populations age, there is an increasing need to optimize endovascular treatment (EVT) for acute ischemic stroke. We harmonized prospective stroke registries from Korea and Taiwan to enable direct comparisons of patient characteristics and clinical outcomes, with a particular focus on the impact of advanced age.

**Methods:** Prospective stroke registries in South Korea (CRCS-K) and Taiwan (TREAT-AIS) were harmonized to form a unified dataset of patients treated with EVT. EVT outcomes included 3-month modified Rankin Scale (mRS), symptomatic intracranial hemorrhage (SICH), and successful recanalization. We assessed the association between age and outcomes in the overall population and in those aged ≥80 years, adjusting for relevant covariates.

**Results:** A total of 9941 EVT cases (7835 from Korea and 2106 from Taiwan) were included. Patients had a mean age of 70.1 ± 12.9 years (57.6% male, median NIHSS: 14 [IQR: 9–19]). Compared to Korean patients, Taiwanese patients had a higher prevalence of vascular risk factors and more severe strokes, contributing to lower rates of favorable 3-month outcomes. Increasing age was associated with poorer EVT results; among patients ≥80 years, only 18% achieved mRS 0–2, compared to 47% of younger patients. However, the risk of SICH did not significantly increase with age (adjusted OR per year: 1.00 [0.99–1.01]; adjusted OR ≥80 years: 1.05 [0.85–1.29]). Pre-stroke functional independence and bridging thrombolysis were identified as key modifiers of better outcomes even in the oldest patients.

**Conclusion:** Taiwanese EVT patients had more risk factors and worse outcomes than Korean patients. Advanced age is associated with poorer functional recovery, yet selected older patients—particularly those who were functionally independent before the stroke or received bridging thrombolysis—demonstrated meaningful benefit. These results underscore the importance of individualized treatment strategies and careful patient selection, especially as populations continue to age.

## Introduction

Timely recanalization is a key determinant of improved outcomes for patients with acute ischemic stroke (AIS).^1^ Robust evidence from multiple randomized trials (RCTs) supports the safety and efficacy of recanalization therapies. In particular, a meta-analysis of five early RCTs revealed that endovascular thrombectomy (EVT) for large vessel occlusion (LVO) reduced disability by a factor of 2.5 compared to standard medical care.^2^ As a result, rapid recanalization strategies – including intravenous thrombolysis and endovascular recanalization – have become integral to AIS management and are now widely adopted in clinical practice.

However, the clinical characteristics and etiologies of AIS differ significantly across ethnic groups. East Asian populations, including those in Korea, Taiwan, China, and Japan, constitute roughly 20% of the global population.^3^ Yet, these groups remain underrepresented in high quality EVT trials.^4^ Notably, patients in East Asia demonstrate a higher incidence of LVO driven by intracranial atherosclerosis (ICAS),^5^ complicating endovascular navigation and increasing the technical complexity of procedures like suction thrombectomy. East Asian patients also have a higher prevalence of hemorrhagic complications post-reperfusion.^6^ These ethnic and regional differences highlight the need for multinational, multicenter registries that focus on specific subpopulations, such as East Asians, to enhance the relevance and generalizability of EVT research.

While advanced age is a well-recognized risk factor for worse outcomes following EVT,^7,8^ the concept of age-related treatment nihilism is unwarranted. Even elderly patients can benefit from EVT, provided that factors like brain frailty, premorbid functional status, and expected clinical outcomes are carefully considered.^9^ As global population age, the incidence of AIS in older adults is expected to rise, making it imperative to understand how age influence EVT outcomes and to develop tailored strategies for older patients.

In this context, researchers in Taiwan and South Korea have established prospective stroke registries – TREAT-AIS in Taiwan and CRCS-K in Korea – to capture real-world EVT practices.^10,11^ To advance understanding of country-specific differences in EVT outcomes across East Asia, these two groups aligned their datasets to form the K-T collaborative EVT registry. In this paper, we describe the organization structure and patient characteristics of this unified registry. Given the rapid aging of both population, we further explore the impact of age on EVT outcomes, providing insights that may guide clinical decision-making in real-world practice.

## Methods

### Establishment and Operation of the K-T Collaborative EVT Registry

The K-T collaborative EVT registry integrates patients-level data from the Clinical Research Collaboration for Stroke in Korea (CRCS-K) and the Taiwan Registry of Endovascular Thrombectomy for Acute Ischemic Stroke (TREAT-AIS), resulting in a harmonized, analyzable dataset. CRCS-K registry, initiated in 2008 and expanding into a broader coverage with prospective collection of stroke outcomes in 2011, had enrolled over 100,000 acute stroke admissions by December 2023.^11^ It includes approximately 700 data elements capturing patient demographics, hyperacute treatment details, time metrics, stroke subtypes, in-hospital management, medications, and event and functional outcomes at 3 months and 1 year. TREAT-AIS, launched in January 2019 with contributions from 10 medical centers and 9 community hospitals in Taiwan, focuses on patients over 20 years old with AIS treated by EVT.^10^ It records stroke severity, arterial occlusion location, recanalization strategies, and 3-month functional recovery.

In mid-2023, researchers from Korea and Taiwan convened in Taipei and agreed to harmonize these registries. For this study, CRCS-K contributed 7835 EVT-treated patients from January 1, 2011 to April 30, 2023, while TREAT-AIS provided 2106 EVT-treated patients enrolled up to April 30, 2023 (Supplemental Table 1). Data dictionaries and coding schemas from both registries were systematically reviewed to identify harmonizable fields. A biostatistician-led team, chaired by Hsieh Y-C, performed data harmonization, culminating in a unified dataset of 9941 EVT patients (7835 from Korea and 2106 from Taiwan) with roughly 200 common data fields.

Ethical approval for analyses was obtained from the Institutional Review Board of the Seoul National University Bundang Hospital (Approval ID, B-2410-928-104). The data that support the findings of this study are available from the corresponding author upon reasonable request.

### Data Collection

Researchers from both countries collaboratively reviewed the source registries’ data dictionaries to identify common variables. These harmonized fields included baseline demographics, pre-stroke functional status, vascular risk factors, use of antithrombotics, stroke subtypes, baseline National Institutes of Health Stroke Scale (NIHSS) scores, LVO location, and key time metrics. The primary EVT outcomes encompassed symptomatic intracerebral hemorrhage (ICH), successful recanalization (modified Thrombolysis in Cerebral Infarction [mTICI] score of 2b–3), 90-day modified Rankin Scale (mRS) scores, and 90-day mortality. Symptomatic ICH was defined as a type 2 parenchymal hematoma on neuroimaging obtained within 24–36 hours post-EVT, accompanied by a ≥4-point increase in the NIHSS.

### Statistical Analyses

We first summarized baseline clinical characteristics and then investigated the association between age and EVT outcomes. Categorical variables were compared using chi-squared tests, parametric variables with independent t-tests, and non-parametric data with median tests. Multivariable logistic regression models, adjusted for clinically relevant covariates, were used to identify independent predictors of outcomes. Non-linearity was examined using restricted cubic splines. A two-tailed P-value <0.05 was considered significant. As this was an exploratory analysis, no corrections for multiple comparisons were applied.^12^ All statistical analyses were performed using R version 4.4.1.

## Results

### Baseline Characteristics and Comparison Between the Two Countries

The K-T collaborative EVT registry comprised 9,941 cases: 7,835 from South Korea (CRCS-K) and 2,106 from Taiwan (TREAT-AIS). As shown in Table 1, the Taiwanese cohort was older on average (71.2 ± 13.3 vs. 69.8 ± 12.8 years; P<0.01) and exhibited a higher prevalence of vascular risk factors including hypertension (73.4% vs. 62.9%), diabetes (34.3% vs. 28.6%), dyslipidemia (51.4% vs. 27.3%), and atrial fibrillation (52.9% vs. 46.6%), while smoking history was more common in the Korean cohort (32.7% vs. 28.1%). TREAT-AIS patients also had lower mean glomerular filtration rates (62.3 ± 31.0 vs. 69.1 ± 32.3) and presented with more severe strokes (median NIHSS: 18 [13–23] vs. 14 [9–18]). Pre-stroke functional independence was less frequent in the Taiwanese group (71.9% vs. 80.5%).

**Table 1.** Overall characteristics of endovascular treatment cases and comparison between South Korea (CRCS-K registry) and Taiwan (TREAT-AIS registry)

|  | Overall | South Korea | Taiwan | P-for-difference |
| --- | --- | --- | --- | --- |
| Number of patients | 9941 | 7835 | 2106 |  |
| Demographic information |  |  |  |  |
| Male sex (%) | 5727 (57.6) | 4547 (58.0) | 1180 ( 56.0) | 0.104 |
| age (mean (SD)) | 70.07 (12.92) | 69.79 (12.78) | 71.13 (13.38) | <0.001 |
| Pre-stroke independence (%) | 7331 (79.1) | 6199 (80.5) | 1132 ( 71.9) | <0.001 |
| Stroke information |  |  |  |  |
| LKW to arrival (hours; median [IQR]) | 2.62 [1.02, 6.38] | 2.62 [1.02, 6.53] | 2.78 [1.03, 5.38] | 0.047 |
| Unwitnessed onset (%) | 3446 (41.6) | 3183 (42.1) | 263 ( 36.9) | 0.008 |
| initial NIHSS score (median [IQR]) | 14 [9- 19] | 14 [9 - 18] | 18 [13 - 23] | <0.001 |
| Bridging thrombolysis (%) | 4297 (43.2) | 3540 (45.2) | 757 ( 35.9) | <0.001 |
| Arrival to IVT (hours; median [IQR]) | 0.60 [0.41, 0.83] | 0.57 [0.40, 0.78] | 0.80 [0.57, 1.00] | <0.001 |
| Arrival to groin puncture (hours; median [IQR]) | 1.83 [1.37, 2.52] | 1.72 [1.32, 2.30] | 2.40 [1.77, 3.37] | <0.001 |
| Ischemic stroke subtype |  |  |  | <0.001 |
| Large artery atherosclerosis (%) | 2738 (27.5) | 2008 (25.6) | 730 ( 34.7) |  |
| Cardioembolism (%) | 4734 (47.6) | 3771 (48.1) | 963 ( 45.7) |  |
| Other-determined etiologies (%) | 387 ( 3.9) | 279 ( 3.6) | 108 ( 5.1) |  |
| Undetermined etiologies (%) | 2082 (20.9) | 1777 (22.7) | 305 ( 14.5) |  |
| Occlusion at posterior circulation (%) | 1339 (13.5) | 1090 (13.9) | 249 ( 11.8) | 0.014 |
| Vascular risk factors |  |  |  |  |
| Hypertension (%) | 6472 (65.1) | 4929 (62.9) | 1543 ( 73.4) | <0.001 |
| Diabetes (%) | 2957 (29.8) | 2237 (28.6) | 720 ( 34.3) | <0.001 |
| Dyslipidemia (%) | 3213 (32.4) | 2136 (27.3) | 1077 ( 51.4) | <0.001 |
| Current smoking (%) | 3152 (31.7) | 2561 (32.7) | 591 ( 28.1) | <0.001 |
| History of stroke (%) | 1888 (19.0) | 1473 (18.8) | 415 ( 19.7) | 0.348 |
| Atrial fibrillation (%) | 4763 (47.9) | 3649 (46.6) | 1114 ( 52.9) | <0.001 |
| History of ischemic heart disease (%) | 1247 (12.5) | 943 (12.0) | 304 ( 14.4) | 0.004 |
| History of heart failure (%) | 351 ( 3.5) | 150 ( 1.9) | 201 ( 9.5) | <0.001 |
| History of BP-lowering medication (%) | 4932 (49.6) | 3970 (50.7) | 962 ( 45.7) | <0.001 |
| History of glucose-lowering medication (%) | 1960 (19.7) | 1584 (20.2) | 376 ( 17.9) | 0.017 |
| History of cholesterol-lowering medication (%) | 704 ( 7.1) | 321 ( 4.1) | 383 ( 18.2) | <0.001 |
| History of antiplatelets use (%) | 2415 (24.3) | 2023 (25.8) | 392 ( 18.6) | <0.001 |
| History of anticoagulants use (%) | 1318 (13.3) | 995 (12.7) | 323 ( 15.3) | 0.002 |
| Concomitant cancer (%) | 519 ( 7.3) | 247 ( 4.9) | 272 ( 12.9) | <0.001 |
| Laboratory information |  |  |  |  |
| Hemoglobin (g/dL; mean (SD)) | 13.4 (2.1) | 13.4 (2.1) | 13.4 (2.3) | 0.375 |
| Platelet count ($\times 10^3$; mean (SD)) | 216 (68) | 217 (66) | 213 (75) | 0.01 |
| PT INR (mean (SD)) | 1.09 (0.38) | 1.08 (0.25) | 1.11 (0.70) | 0.001 |
| Hemoglobin A1c (%; mean (SD)) | 6.2 (1.2) | 6.1 (1.1) | 6.3 (1.4) | <0.001 |
| Glucose level on arrival (mg/dL; mean (SD)) | 142 (53) | 141 (52) | 146 (58) | <0.001 |
| Blood urea nitrogen (mg/dL; mean (SD)) | 18.2 (9.0) | 17.9 (8.0) | 19.7 (12.4) | <0.001 |
| Creatinine (mg/dL; mean (SD)) | 1.03 (0.81) | 0.99 (0.70) | 1.19 (1.12) | <0.001 |
| Glucose level, fasting (mg/dL; mean (SD)) | 133 (51) | 131 (50) | 142 (55) | <0.001 |
| Total cholesterol (mg/dL; mean (SD)) | 161 (42) | 162 (42) | 157 (42) | <0.001 |
| estimated glomerular filtration rate (mean (SD)) | 67.7 (32.2) | 69.1 (32.3) | 62.3 (31.0) | <0.001 |
| Treatment outcomes |  |  |  |  |
| mRS score 0 - 1 at 3 months (%) | 2421 (27.1) | 2055 (28.2) | 366 ( 22.2) | <0.001 |
| mRS score 0 - 2 at 3 months (%) | 3569 (40.0) | 2996 (41.1) | 573 ( 34.7) | <0.001 |
| Mortality up to 3 months (%) | 1138 (12.7) | 1036 (14.2) | 102 ( 6.2) | <0.001 |
| Post-EVT TICI 2b-3 (%) | 6562 (81.1) | 4793 (80.0) | 1769 ( 84.0) | <0.001 |
| Symptomatic hemorrhage, post-EVT (%) | 730 ( 7.3) | 621 ( 7.9) | 109 ( 5.2) | <0.001 |

P-values from the comparison between Korean and Taiwanese EVT cases

Treatment timelines and patterns varied. Time from last known well to ER arrival was slightly longer in TREAT-AIS (medians [interquartile ranges]; 2.78 [1.03 – 5.38] vs. 2.62 [1.02 – 6.53] hours), resulting in lower IV thrombolysis rates (35.9% vs. 45.2%). Similarly, arrival-to-groin puncture times differed between cohorts (2.40 [1.77–3.37] hours in TREAT-AIS vs. 1.72 [1.32–2.30] hours in CRCS-K).

### Prognostic Factors for Outcomes After Endovascular Recanalization Treatment

Successful recanalization rates were similar (84.0% vs. 80.0%), as were symptomatic hemorrhagic transformation rates (5.2% vs. 7.9%). However, functional independence at 90 days was less common in TREAT-AIS (34.7% vs. 41.1%), while 90-day mortality was higher in CRCS-K (14.2% vs. 6.2%). Multivariable logistic regression identified key predictors of outcomes (Supplemental Table 2). Age, pre-stroke functional independence, baseline NIHSS, diabetes, initial hemoglobin, glucose, blood urea nitrogen levels, posterior circulation occlusion, and bridging thrombolysis were each associated with functional recovery and mortality. For SICH, age, pre-stroke independence, baseline NIHSS, anterior circulation occlusion, and platelet and glucose levels, while age, sex, pre-stroke independence, and stroke subtype predicted successful recanalization.

### Detailed Analyses of Age as a Prognosticator After EVT

The mean age of EVT-treated patients was 70.1 ± 12.9 years (range: 7–103). Of these, 24.9% were ≥80 years old, and 3.0% were ≥90 years old. (Supplemental Figure 1). Elderly patients (≥80 years) were more likely to be from Taiwan, were less frequently male, and less often functionally independent prior to stroke. They had more severe neurological deficits at presentation, arrived later, and were less likely to receive IV thrombolysis (34.6% vs. 46.1%). Compared to younger counterparts, older patients had lower rates of favorable functional outcomes (18.1% vs. 46.9%), higher mortality (19.7% vs. 10.5%; Figure 1), and lower successful recanalization rates (78.6% vs. 81.9%; Table 2, Supplemental Table 3).

**Table 2.**
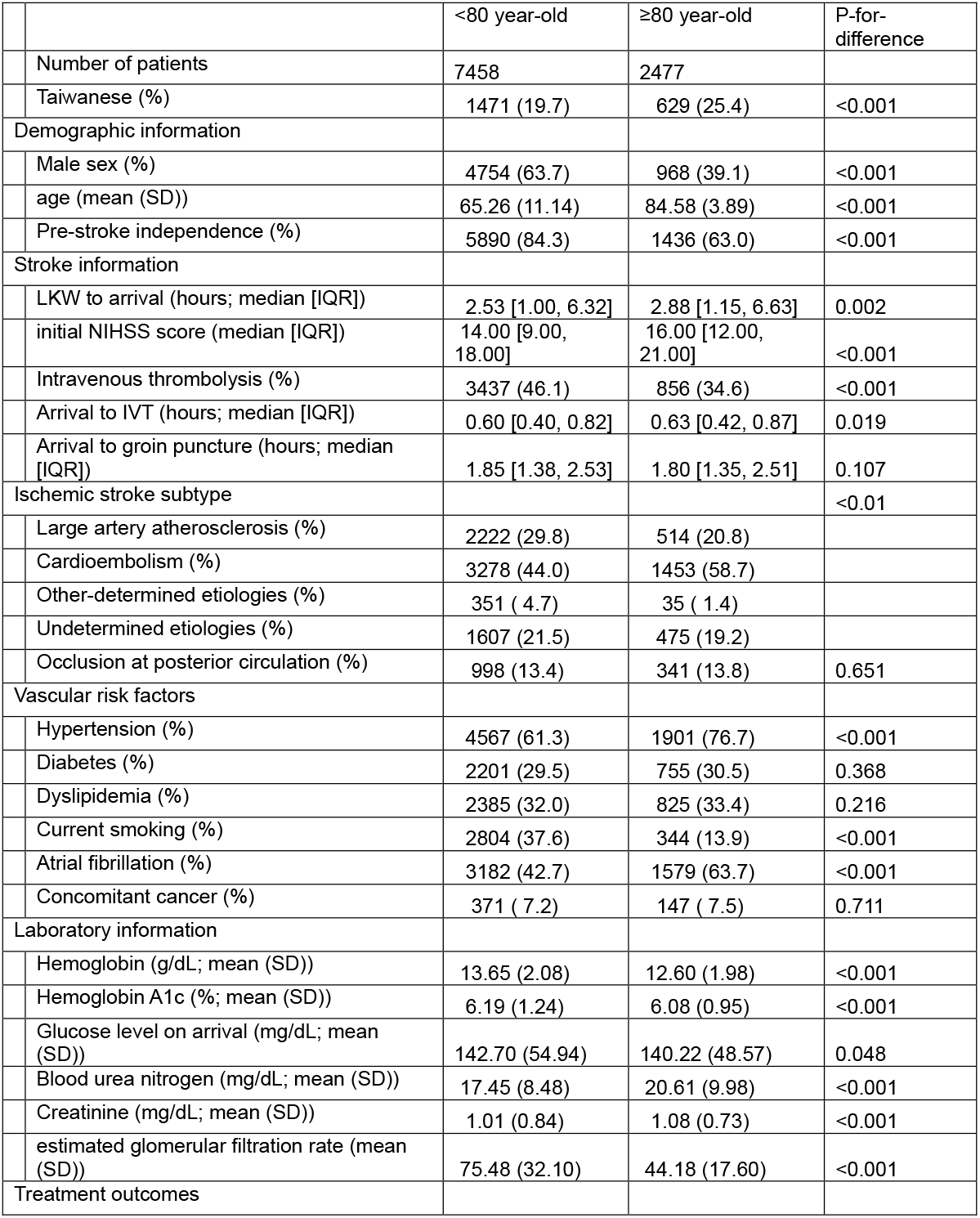
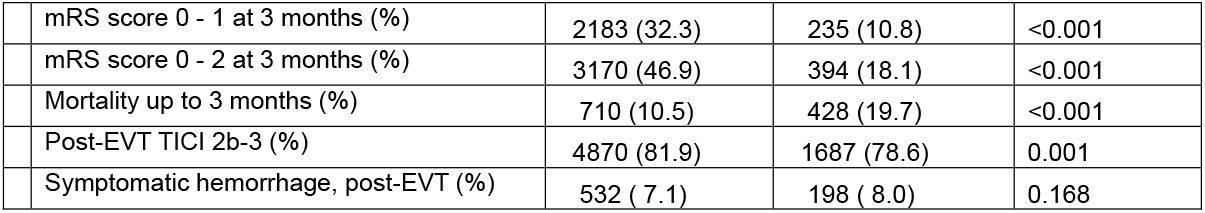
Selected baseline characteristics comparing EVT patients with <80 year-old and ≥80 year-old.

|  | <80 year-old | ≥80 year-old | P-for-difference |
| --- | --- | --- | --- |
| Number of patients | 7458 | 2477 |  |
| Taiwanese (%) | 1471 (19.7) | 629 (25.4) | <0.001 |
| Demographic information |  |  |  |
| Male sex (%) | 4754 (63.7) | 968 (39.1) | <0.001 |
| age (mean (SD)) | 65.26 (11.14) | 84.58 (3.89) | <0.001 |
| Pre-stroke independence (%) | 5890 (84.3) | 1436 (63.0) | <0.001 |
| Stroke information |  |  |  |
| LKW to arrival (hours; median [IQR]) | 2.53 [1.00, 6.32] | 2.88 [1.15, 6.63] | 0.002 |
| initial NIHSS score (median [IQR]) | 14.00 [9.00, 18.00] | 16.00 [12.00, 21.00] | <0.001 |
| Intravenous thrombolysis (%) | 3437 (46.1) | 856 (34.6) | <0.001 |
| Arrival to IVT (hours; median [IQR]) | 0.60 [0.40, 0.82] | 0.63 [0.42, 0.87] | 0.019 |
| Arrival to groin puncture (hours; median [IQR]) | 1.85 [1.38, 2.53] | 1.80 [1.35, 2.51] | 0.107 |
| Ischemic stroke subtype |  |  | <0.01 |
| Large artery atherosclerosis (%) | 2222 (29.8) | 514 (20.8) |  |
| Cardioembolism (%) | 3278 (44.0) | 1453 (58.7) |  |
| Other-determined etiologies (%) | 351 ( 4.7) | 35 ( 1.4) |  |
| Undetermined etiologies (%) | 1607 (21.5) | 475 (19.2) |  |
| Occlusion at posterior circulation (%) | 998 (13.4) | 341 (13.8) | 0.651 |
| Vascular risk factors |  |  |  |
| Hypertension (%) | 4567 (61.3) | 1901 (76.7) | <0.001 |
| Diabetes (%) | 2201 (29.5) | 755 (30.5) | 0.368 |
| Dyslipidemia (%) | 2385 (32.0) | 825 (33.4) | 0.216 |
| Current smoking (%) | 2804 (37.6) | 344 (13.9) | <0.001 |
| Atrial fibrillation (%) | 3182 (42.7) | 1579 (63.7) | <0.001 |
| Concomitant cancer (%) | 371 ( 7.2) | 147 ( 7.5) | 0.711 |
| Laboratory information |  |  |  |
| Hemoglobin (g/dL; mean (SD)) | 13.65 (2.08) | 12.60 (1.98) | <0.001 |
| Hemoglobin A1c (%; mean (SD)) | 6.19 (1.24) | 6.08 (0.95) | <0.001 |
| Glucose level on arrival (mg/dL; mean (SD)) | 142.70 (54.94) | 140.22 (48.57) | 0.048 |
| Blood urea nitrogen (mg/dL; mean (SD)) | 17.45 (8.48) | 20.61 (9.98) | <0.001 |
| Creatinine (mg/dL; mean (SD)) | 1.01 (0.84) | 1.08 (0.73) | <0.001 |
| estimated glomerular filtration rate (mean (SD)) | 75.48 (32.10) | 44.18 (17.60) | <0.001 |
| Treatment outcomes |  |  |  |
| mRS score 0 - 1 at 3 months (%) | 2183 (32.3) | 235 (10.8) | <0.001 |
| mRS score 0 - 2 at 3 months (%) | 3170 (46.9) | 394 (18.1) | <0.001 |
| Mortality up to 3 months (%) | 710 (10.5) | 428 (19.7) | <0.001 |
| Post-EVT TICI 2b-3 (%) | 4870 (81.9) | 1687 (78.6) | 0.001 |
| Symptomatic hemorrhage, post-EVT (%) | 532 ( 7.1) | 198 ( 8.0) | 0.168 |

**Figure 1.**
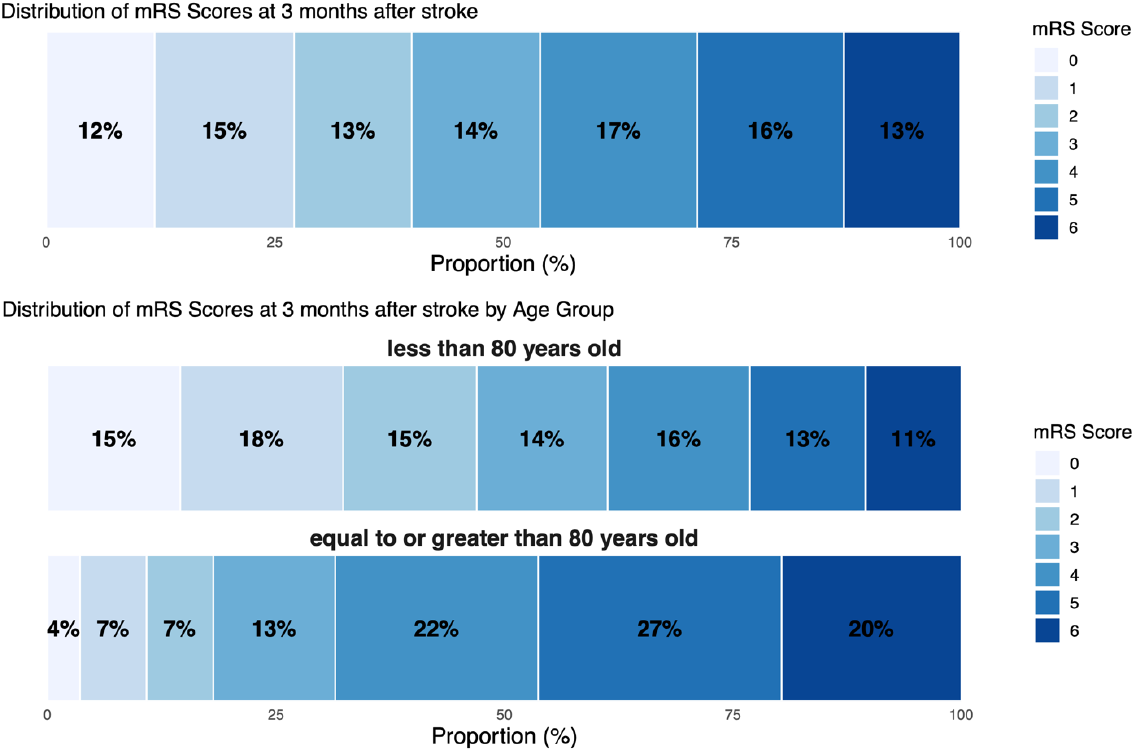
Distribution of mRS scores at 3 months after stroke

Multivariable modeling underscored the influence of age on outcomes, with adjusted ORs of 0.35 [0.30–0.40] for mRS 0–2, 1.49 [1.25–1.76] for mortality, and 0.75 [0.64–0.87] for TICI 2b–3. In contrast, the incidence of SICH was comparable across age groups (7.1% vs. 7.3%, OR: 1.04 [0.84–1.28]; Table 3). Age exhibited a nonlinear relationship with functional recovery and mortality, most pronounced between 75–85 years (Figure 2; OR per 1-year increase for mRS 0–2: 0.90 [0.87–0.93]; OR for mortality: 1.05 [1.01–1.10]), while its relationship with SICH and successful recanalization appeared more linear (Supplemental Table 4). Stratified analyses consistently identified pre-stroke independence and baseline NIHSS as key modifiers of outcomes across all ages, while baseline glucose and cardioembolic stroke etiology remained influential for SICH and recanalization, respectively (Supplemental Tables 5– 8).

**Table 3.**
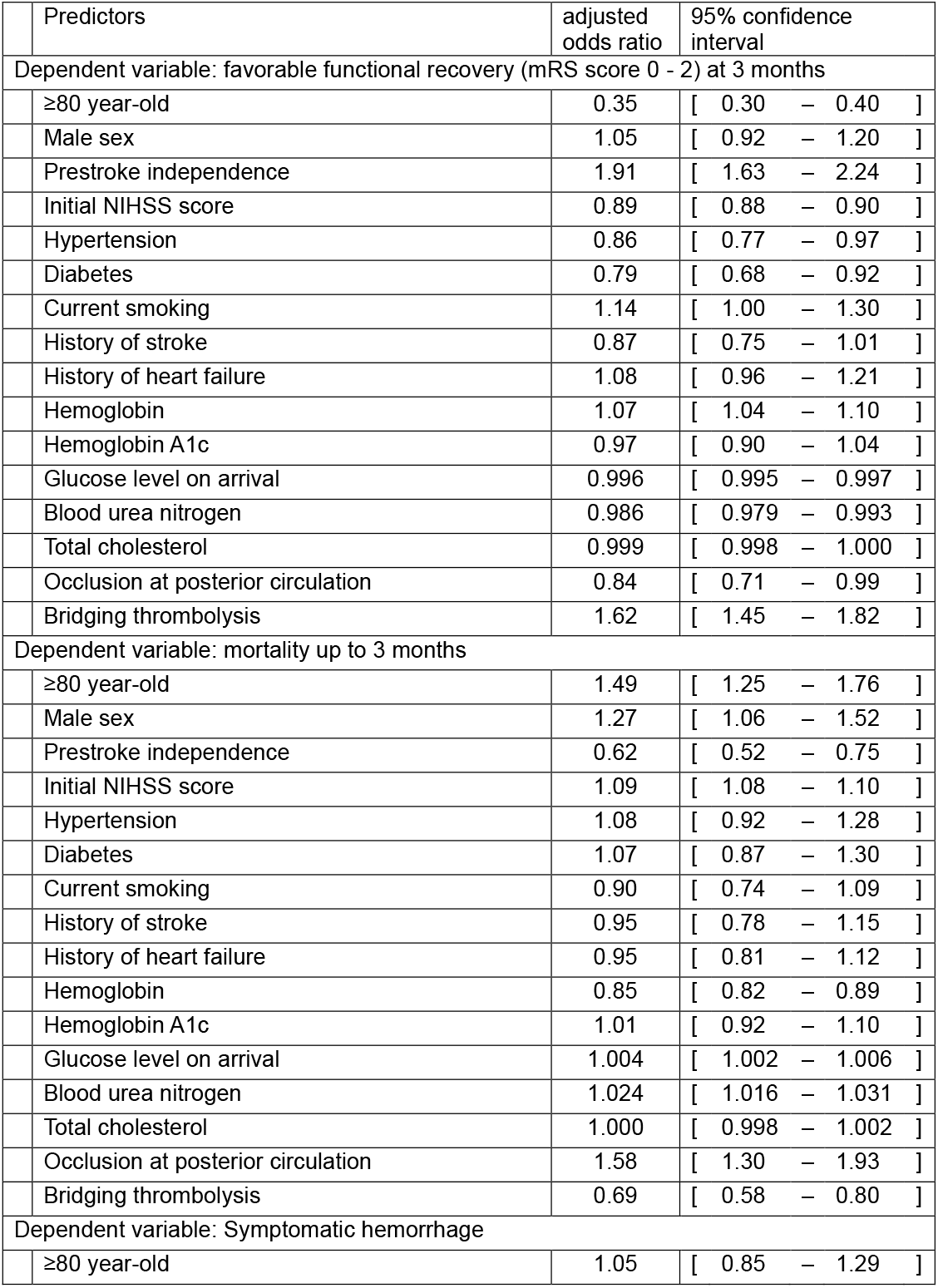
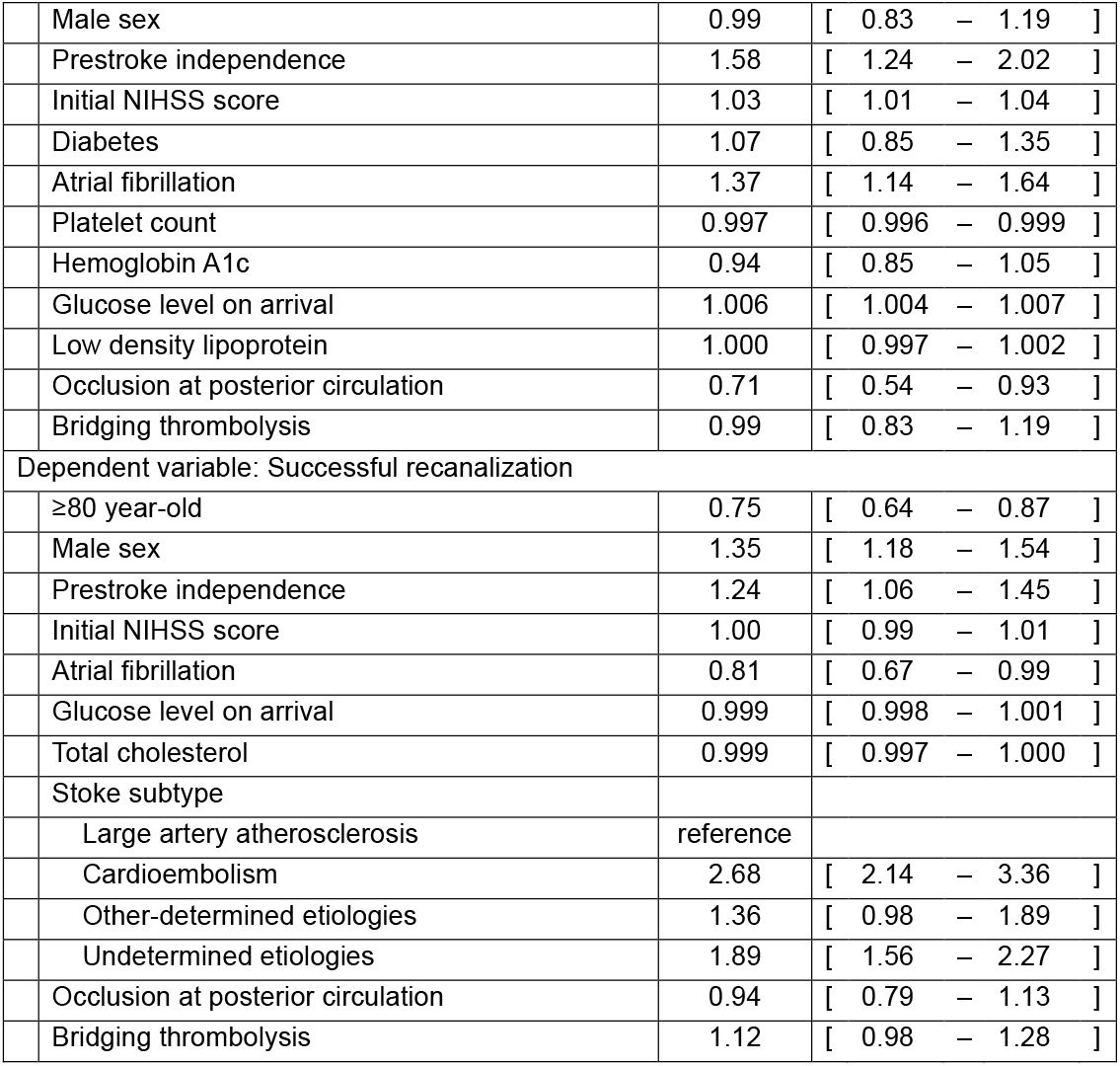
Effect of age strata (< 80 versus ≥80 years old) on the EVT outcomes.

| Predictors | adjusted odds ratio | 95% confidence interval |
| --- | --- | --- |
| Dependent variable: favorable functional recovery (mRS score 0 - 2) at 3 months |  |  |
| ≥80 year-old | 0.35 | [ 0.30 – 0.40 ] |
| Male sex | 1.05 | [ 0.92 – 1.20 ] |
| Prestroke independence | 1.91 | [ 1.63 – 2.24 ] |
| Initial NIHSS score | 0.89 | [ 0.88 – 0.90 ] |
| Hypertension | 0.86 | [ 0.77 – 0.97 ] |
| Diabetes | 0.79 | [ 0.68 – 0.92 ] |
| Current smoking | 1.14 | [ 1.00 – 1.30 ] |
| History of stroke | 0.87 | [ 0.75 – 1.01 ] |
| History of heart failure | 1.08 | [ 0.96 – 1.21 ] |
| Hemoglobin | 1.07 | [ 1.04 – 1.10 ] |
| Hemoglobin A1c | 0.97 | [ 0.90 – 1.04 ] |
| Glucose level on arrival | 0.996 | [ 0.995 – 0.997 ] |
| Blood urea nitrogen | 0.986 | [ 0.979 – 0.993 ] |
| Total cholesterol | 0.999 | [ 0.998 – 1.000 ] |
| Occlusion at posterior circulation | 0.84 | [ 0.71 – 0.99 ] |
| Bridging thrombolysis | 1.62 | [ 1.45 – 1.82 ] |
| Dependent variable: mortality up to 3 months |  |  |
| ≥80 year-old | 1.49 | [ 1.25 – 1.76 ] |
| Male sex | 1.27 | [ 1.06 – 1.52 ] |
| Prestroke independence | 0.62 | [ 0.52 – 0.75 ] |
| Initial NIHSS score | 1.09 | [ 1.08 – 1.10 ] |
| Hypertension | 1.08 | [ 0.92 – 1.28 ] |
| Diabetes | 1.07 | [ 0.87 – 1.30 ] |
| Current smoking | 0.90 | [ 0.74 – 1.09 ] |
| History of stroke | 0.95 | [ 0.78 – 1.15 ] |
| History of heart failure | 0.95 | [ 0.81 – 1.12 ] |
| Hemoglobin | 0.85 | [ 0.82 – 0.89 ] |
| Hemoglobin A1c | 1.01 | [ 0.92 – 1.10 ] |
| Glucose level on arrival | 1.004 | [ 1.002 – 1.006 ] |
| Blood urea nitrogen | 1.024 | [ 1.016 – 1.031 ] |
| Total cholesterol | 1.000 | [ 0.998 – 1.002 ] |
| Occlusion at posterior circulation | 1.58 | [ 1.30 – 1.93 ] |
| Bridging thrombolysis | 0.69 | [ 0.58 – 0.80 ] |
| Dependent variable: Symptomatic hemorrhage |  |  |
| ≥80 year-old | 1.05 | [ 0.85 – 1.29 ] |

|  |  |  |  |
| --- | --- | --- | --- |
|  | Male sex | 0.99 | [ 0.83 – 1.19 ] |
|  | Prestroke independence | 1.58 | [ 1.24 – 2.02 ] |
|  | Initial NIHSS score | 1.03 | [ 1.01 – 1.04 ] |
|  | Diabetes | 1.07 | [ 0.85 – 1.35 ] |
|  | Atrial fibrillation | 1.37 | [ 1.14 – 1.64 ] |
|  | Platelet count | 0.997 | [ 0.996 – 0.999 ] |
|  | Hemoglobin A1c | 0.94 | [ 0.85 – 1.05 ] |
|  | Glucose level on arrival | 1.006 | [ 1.004 – 1.007 ] |
|  | Low density lipoprotein | 1.000 | [ 0.997 – 1.002 ] |
|  | Occlusion at posterior circulation | 0.71 | [ 0.54 – 0.93 ] |
|  | Bridging thrombolysis | 0.99 | [ 0.83 – 1.19 ] |
| Dependent variable: Successful recanalization |  |  |  |
|  | ≥80 year-old | 0.75 | [ 0.64 – 0.87 ] |
|  | Male sex | 1.35 | [ 1.18 – 1.54 ] |
|  | Prestroke independence | 1.24 | [ 1.06 – 1.45 ] |
|  | Initial NIHSS score | 1.00 | [ 0.99 – 1.01 ] |
|  | Atrial fibrillation | 0.81 | [ 0.67 – 0.99 ] |
|  | Glucose level on arrival | 0.999 | [ 0.998 – 1.001 ] |
|  | Total cholesterol | 0.999 | [ 0.997 – 1.000 ] |
|  | Stoke subtype |  |  |
|  | Large artery atherosclerosis | reference |  |
|  | Cardioembolism | 2.68 | [ 2.14 – 3.36 ] |
|  | Other-determined etiologies | 1.36 | [ 0.98 – 1.89 ] |
|  | Undetermined etiologies | 1.89 | [ 1.56 – 2.27 ] |
|  | Occlusion at posterior circulation | 0.94 | [ 0.79 – 1.13 ] |
|  | Bridging thrombolysis | 1.12 | [ 0.98 – 1.28 ] |

**Figure 2.**
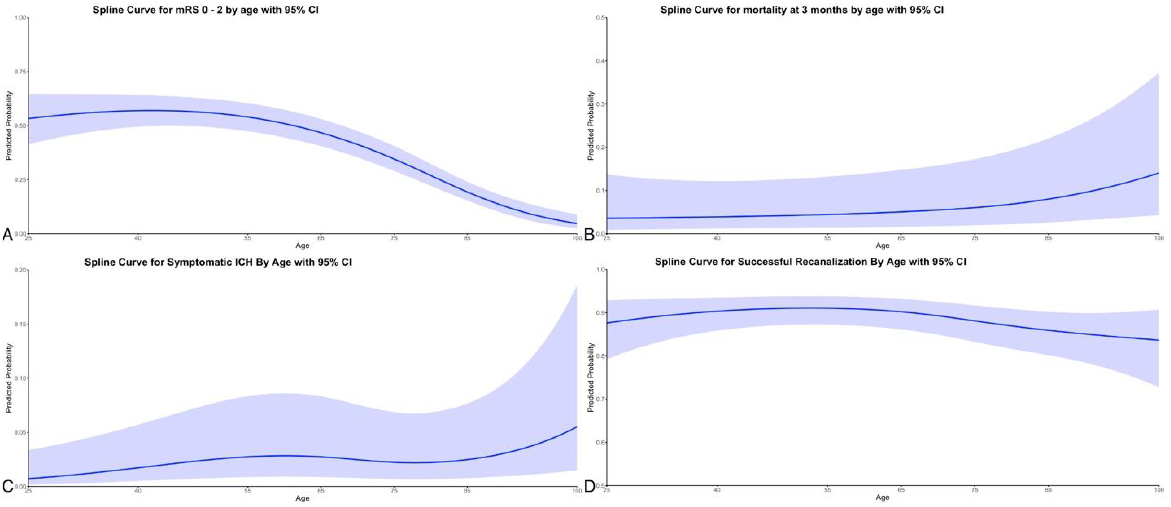
Non-linear association between age and selected outcomes following endovascular treatment (EVT) (A) Favorable functional recovery, defined as a modified Rankin Scale (mRS) score of 0-2 at 3 months, (B) Mortality within 3 months post-stroke, (C) Incidence of symptomatic intracerebral hemorrhage, and (D) Successful recanalization, defined as post-EVT TICI 2b-3. These associations were modeled using restricted cubic spline curves (dark blue lines) with 95% confidence intervals (light blue shading).

In patients ≥80 years, favorable recovery was predicted by younger age within this subgroup, female sex, pre-stroke independence, lower baseline NIHSS, and bridging thrombolysis. Mortality predictors included pre-stroke independence, pre-EVT tPA use, initial hemoglobin, eGFR, and bridging thrombolysis (Table 4).

**Table 4.** Prognostic factors for functional recovery and mortality after endovascular treatment in ≥80 year-old patients.

| Predictors | adjusted odds ratio | 95% confidence interval |
| --- | --- | --- |
| Dependent variable: favorable functional recovery (mRS score 0 - 2) at 3 months |  |  |
| Age | 0.93 | [ 0.90 – 0.97 ] |
| Male sex | 1.44 | [ 1.10 – 1.90 ] |
| Prestroke independence | 2.05 | [ 1.51 – 2.78 ] |
| Initial NIHSS score | 0.90 | [ 0.88 – 0.92 ] |
| Diabetes | 0.75 | [ 0.55 – 1.03 ] |
| Current smoking | 1.13 | [ 0.79 – 1.61 ] |
| History of stroke | 0.82 | [ 0.59 – 1.14 ] |
| Hemoglobin | 1.04 | [ 0.97 – 1.11 ] |
| Initial glucose | 0.998 | [ 0.994 – 1.001 ] |
| Estimated GFR | 0.997 | [ 0.989 – 1.004 ] |
| Bridging thrombolysis | 1.62 | [ 1.24 – 2.10 ] |
| Dependent variable: mortality up to 3 months |  |  |
| Age | 1.04 | [ 1.003 – 1.07 ] |
| Male sex | 1.59 | [ 1.24 – 2.05 ] |
| Prestroke independence | 0.76 | [ 0.59 – 0.97 ] |
| Initial NIHSS score | 1.07 | [ 1.05 – 1.09 ] |
| History of ischemic heart disease | 1.27 | [ 0.93 – 1.74 ] |
| History of heart failure | 2.73 | [ 1.56 – 4.79 ] |
| Hemoglobin | 0.90 | [ 0.84 – 0.95 ] |
| Prothrombin INR | 1.48 | [ 0.92 – 2.36 ] |
| Initial glucose | 1.004 | [ 1.002 – 1.006 ] |
| Estimated GFR | 0.988 | [ 0.980 – 0.996 ] |
| Posterior circulation occlusion | 1.19 | [ 0.86 – 1.65 ] |
| Bridging thrombolysis | 0.74 | [ 0.57 – 0.96 ] |

## Discussion

By integrating two national stroke registries from Korea and Taiwan, we created a harmonized dataset of 9,941 EVT-treated patients. Our analyses revealed that Taiwanese patients tended to have more complex vascular risk profiles and more severe neurological deficits, correlating with lower rates of functional recovery. We also confirmed that age strongly influences EVT outcomes, particularly between 75 and 85 years. Notably, however, the risk of symptomatic hemorrhagic transformation did not differ significantly by age.

This multinational registry provides unique insights into EVT performance in East Asia. While major RCTs have firmly established EVT’s efficacy,^13^ challenges remain in translating these findings to diverse populations and in optimizing patient selection.^14^ The inclusion of patients who may be underrepresented in clinical trials—such as the very elderly or those with extensive comorbidities and East Asians—enables a more nuanced understanding of EVT’s real-world impact and supports more individualized treatment approaches.

Our results highlight how differences in baseline demographics, risk factors, and care pathways between Taiwan and Korea can shape EVT outcomes. Multinational datasets are instrumental for identifying such variations, informing clinicians, and guiding policymakers to refine care models.^15^ More granular insights into factors like stroke subtype, bridging therapies, comorbidities and baseline imaging can help tailor EVT strategies to different healthcare contexts and population needs.

Consistent with previous studies, we found that advanced age often portends worse functional recovery and higher mortality.^7,8^ In our dataset, patients aged 80 and older had significantly worse outcomes compared to younger patients, with lower rates of functional independence (10.8% vs. 32.3%) and higher mortality (19.7% vs. 10.5%). These results align with previous studies that identified age as a significant predictor of poor outcomes and futile revascularization after EVT.^16,17^ When compared to a post-hoc analysis of HERMES collaboration, ≥80 year-old patients from Korea and Taiwan achieved comparable functional recovery with higher mortality and SICH.^18^

Several factors likely contribute to the worse outcomes observed in older patients. Older patients tend to have a higher burden of comorbidities, more severe strokes at presentation, and delayed treatment initiation, all of which compromise functional gains.^19,20^ Yet, our findings, consistent with previous studies, do not support an age-based exclusion from EVT.^21,22^ Even among the elderly, approximately 16% achieved favorable outcomes. Moreover, pre-stroke functional independence and the use of bridging IV thrombolysis were strong predictors of improved outcomes in the elderly. This underscores the importance of considering factors beyond chronological age–such as ischemic core and penumbra volumes, stroke location, comorbidities, frailty, and bridging thrombolysis–in patient selection.^23^

The strengths of our study lie in its large scale, multinational design, and prospective data collection. By analyzing a broad range of clinical profiles, we offer insights that may be more generalizable than single-country or strictly trial-based data.^24^ Nonetheless, this analysis has limitations. Differences in data collection methods and definitions between registries may affect comparability. The inclusion periods vary by registry, and detailed imaging data or post-discharge care information were not available. Such factors, along with the retrospective nature of these analyses, warrant cautious interpretation.

In conclusion, our multinational approach demonstrates the value of harmonizing stroke registries to advance global stroke care. We show that while advanced age is associated with poorer EVT outcomes, carefully selected elderly patients can still derive meaningful benefit. Future collaborative efforts should focus on refining EVT techniques, integrating imaging and longitudinal follow-up data, and systematically addressing regional variations in care delivery. Such endeavors will ultimately improve EVT strategies and outcomes across East Asia, ensuring that an ever-widening population of patients receives the most effective acute stroke interventions.

## Data Availability

The data that support the findings of this study are available from the corresponding author upon reasonable request.

## Acknowledgement

A complete list of participating researchers is provided as an Appendix in the Supplemental Data.

